# Real burden of mucormycosis diagnosed over five years in a French medical center : a retrospective study

**DOI:** 10.1101/2025.10.30.25338085

**Authors:** Hugues Beudet, Jacques Sevestre, Hugo Bes Berlandier, Léa Delorme, Sebastien Cortaredona, Pierre Berger, Jean-Christophe Lagier, Stéphane Ranque, Nadim Cassir, Pierre Dudouet

**Author notes:** **Corresponding author:**, IHU - Méditerranée Infection, 19-21 boulevard Jean Moulin, 13005 Marseille, France.

## Abstract

Mucormycosis, a rare but fatal invasive fungal infection, affects immunocompromised patients. Despite recent advancements in diagnostic tools and treatments, mortality rates remain high, reaching 79% at ninety days in disseminated forms. Quantitative Polymerase Chain Reaction (qPCR) based methods are now widely available, and help to discern colonisation and infection, although this remains a real challenge. This study aims to describe the epidemiological and clinical characteristics of a French cohort of patients with presence of Mucorales in clinical samples. With a retrospective monocentric study from 2017 to 2022, we investigated risk factors associated with proven or probable mucormycosis and in-hospital mortality. Patients included had at least one Mucorale-positive culture, or positive PCR assay detecting Mucorales. Clinical, microbiological, and hospital management data were collected and analyzed using univariate and multivariable statistical models. Among our 85 identified patients, in-hospital mortality was significantly associated with positive cultures obtained from deep respiratory samples (OR=16.6, p=0.017) and diagnosis based on EORTC+PCR criteria (OR=12.4, p=0.016). Principal-component analysis revealed a homogeneous group of patients with positive deep respiratory swabs, haemopathy, ICU admission and a greater death ratio. We completed with an expert analyses on diagnosis, which revealed diagnostic variability with an overall Krippendorff’s alpha about 0.32. Mucormycosis remains a challenging condition due to its rarity, severity, and diagnostic limitations. Incorporating PCR into diagnostic criteria and implementing early and tailored management for at-risk patients may improve clinical outcomes. Prospective studies are required to validate these findings and refine management strategies for this life-threatening infection.

## INTRODUCTION

Mucormycosis is a severe invasive mold related infection and has recently been included in the WHO list of high priority pathogens. It is responsible for the highest mortality rate in Europe as an invasive fungal infection, and its incidence has been increasing over the past decades (1,2). Patients at risk include the immunocompromised, such as those who received allogenic hematopoietic system cell transplantation (BMT), solid organ transplantation (SOT), severe neutropenia, but also those presenting with diabetes mellitus and suffering traumatic wounds (3,4). Different clinical presentations have been observed depending on the predisposing clinical condition and geographical distribution. In Europe, pulmonary and disseminated mucormycosis are more commonly reported in immunocompromised patients: SOT and haematological malignancies (HM), whereas COVID-19-associated or rhino-cerebral damage are described in India with diabetes mellitus as the underlying condition (4,5). Maximising survival rates requires rapid diagnosis and therapy, including a combination of liposomal amphotericin B and early extensive surgical treatment where possible (6,7).

Although there has been a marked improvement over the last decade, the prognosis and diagnostic sensitivity remain poor. For example, in French cohorts, the 90-day mortality rates for pulmonary and disseminated mucormycosis were 59% and 79%, respectively (8,9). Since the definition of invasive fungal disease from the 2019 European Organisation for Research and Treatment of Cancer/Mycoses Study Group (EORTC/MSG), new tools and awareness have been developed, allowing for more accurate diagnosis and reducing the time taken to diagnose the disease (10,11). In particular, quantitative polymerase chain reaction (qPCR)-based methods for diagnosing mucormycosis are now widely available in France at reasonable cost, enabling them to be used as a routine technique (10,12–14). A new challenge will be to determine the appropriate role of these new assays in the decision-making process, which may require adjustments to existing recommendations. Mucormycosis remains a rare infection and has been described in literature that is limited by a small patient sample. Recently, there has been unprecedented interest in this previously neglected infection, with several studies reporting larger patient numbers (3,8).

Our study therefore aims to describe a cohort of patients presenting positive Mucorales samples at a French medical centre, assessing factors associated with proven or probable mucormycosis and mortality, and providing resources to help manage these infections.

## PATIENTS/MATERIALS AND METHODS

We conducted a retrospective observational epidemiological study, from 01/01/2017 to 31/12/2022, with university mycology laboratory’s data which centralises requests for the microbiological diagnosis of mucormycosis from 3 university hospital centers and one cancer research centre in Marseille, south of France. Patients were included if they presented with at least one positive Mucorales sample result in our laboratory. No exclusion criteria were applied (Figure 1).

**Figure 1.**
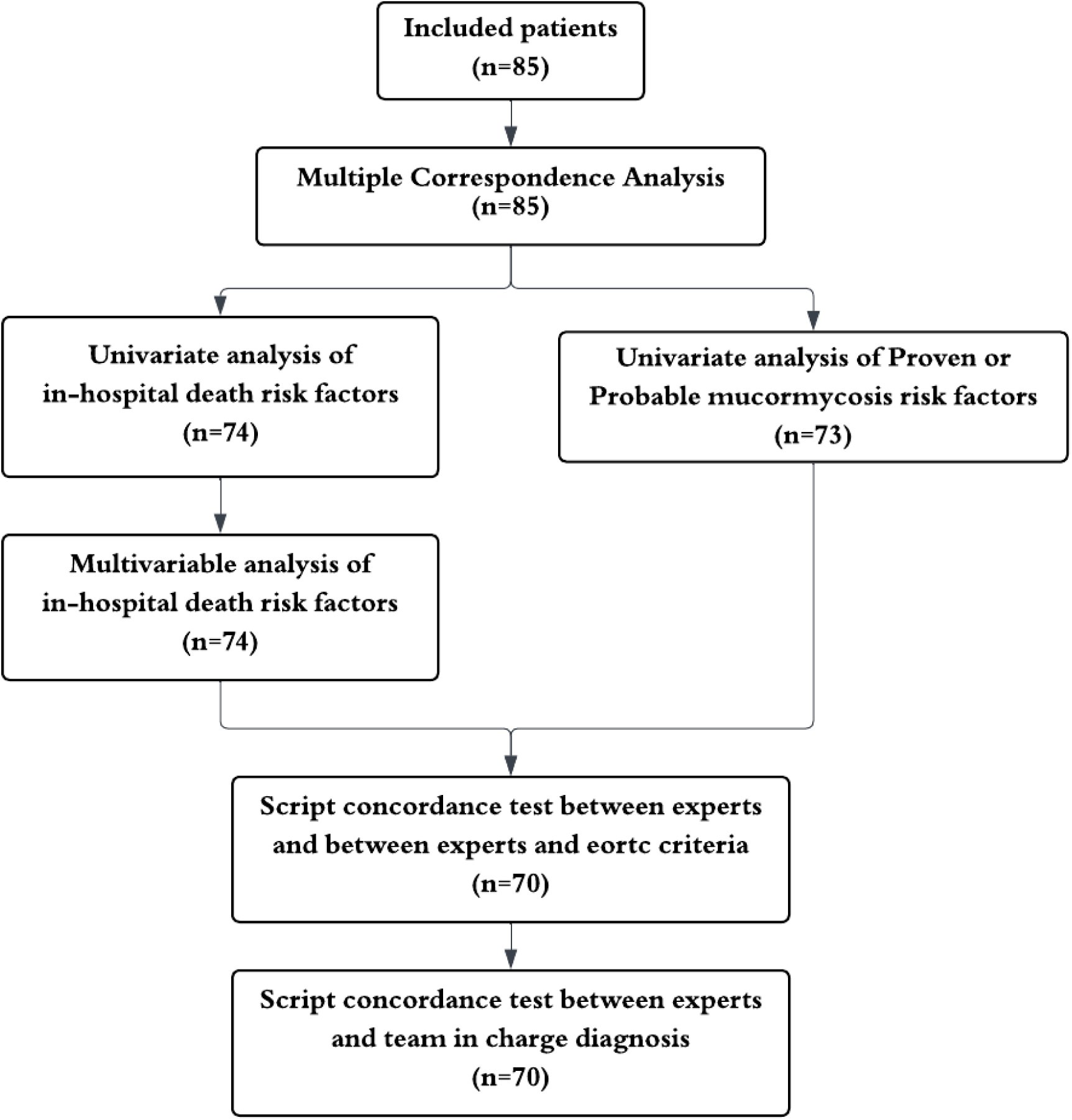
Flow Chart.

### Collected variable of interest

#### Clinical background

The patients’ clinical backgrounds were collected. Immunocompromised patients were classified as having HM, active cancer, BMT or SOT. Based on literature data, the presence of uncontrolled diabetes, traumatic wounds, or burns was investigated (3,5,15,16).

Then, patient clinical evolution, intensive care admission, length of hospitalisation, diagnosis delay, recourse to medical or surgical treatment, and in-hospital death, were all collected to assess the disease management. Patients were categorised as having suspected or ruled-out mucormycosis, depending on whether they received treatment from the medical team in charge. Treatment duration corresponded to the day of treatment against mucormycosis initiation until the end of treatment or the last reported follow-up. In-hospital mortality was recorded if the patient died before discharge. The date of diagnosis corresponded to the first microbiological evidence of mucormycosis. To assess the time taken for each microbiological diagnostic tool, we recorded the time elapsed between sample collection and communication of results to clinicians.

#### Complementary examinations

Data obtained from biological and microbiological assays were collected. Neutropenia was defined as having a white blood cell count below 1,500 G/l within one month of the diagnosis of infection. Only cultures were used to identify the species. PCR testing for Mucorales was only available for diagnosis from 2020 to 2022. Based on previous data showing that 36% of cases of aspergillosis co-infection occur in pulmonary mucormycosis (8) and the recent increase in incidence of mucormycosis associated with SARS-CoV-2 infection (17), the presence of other viral or fungal infections was screened for. It was and recorded if they occurred within one month of mucormycosis diagnosis, regardless of whether they were tested positive in the same anatomical aera. Histology results (assessing tissue invasion) and radiological explorations (e.g. CT or MRI scans) were recorded whenever findings were compatible with a diagnosis of mucormycosis (for example, showing inverted halo patterns) (8).

### Laboratory protocols and technics

Upon sampling, specimens were addressed to the microbiology laboratory for diagnostic analyses. For ENT biopsies, specimens were assessed by direct examination, culture, and PCR analysis. Direct examination was performed over dry smears stained with MYCETBLU (Biosynex, Strasbourg, France). Cultures were inoculated on Blood agar, Chocolate agar and Sabouraud Dextrose agar (BioMérieux, Marcy-L’Etoile, France). Biopsy specimens also underwent digestion for PCR analyses targeting Aspergillus spp. and Mucorales using the MycoGenie kit (AdemTech, Pessac, France). When a mold was grown, species level identification used Matrix-Assisted Laser Desorption-Ionization/Time-Of-Flight Mass Spectrometry (MALDI-TOF MS) via a MicroFlex automaton (Bruker, Bremen, Germany) using an in-house reference spectra library. Before February 2022, Aspergillus was detected via an in-house qPCR assay targeting the 18S rDNA gene, and Mucorales via in-house assays targeting the genera Absidia, Mucor and Rhizomucor. Respiratory samples were assessed by culture and PCR analyses. Culture methods included inoculation over Chocolate agar, Nalidixic acid/Colistin blood agar, MacConkey agar and Sabouraud Dextrose agar (BioMérieux, Marcy-L’Etoile, France). Specimens also underwent digestion for PCR analyses targeting Aspergillus spp. and Mucorales using the MycoGenie kit (AdemTech, Pessac, France). When a mold was grown, species level identification used Matrix-Assisted Laser Desorption-Ionization/Time-Of-Flight Mass Spectrometry (MALDI-TOF MS) via a MicroFlex automaton (Bruker, Bremen, Germany) using an in-house reference spectra library. Before February 2022, Aspergillus was detected via an in-house qPCR assay targeting the 18S rDNA gene, and Mucorales via in-house assays targeting the genera Absidia, Mucor and Rhizomucor. Furthermore, detection of Mucorales circulating DNA in blood was performed on peripheral blood collected over EDTA-coated sampling tubes, and underwent digestion for PCR analyses targeting Mucorales using the MycoGenie kit (AdemTech, Pessac, France). Before February 2022, detection of Mucorales DNA was assessed via in-house assays targeting the genera Absidia, Mucor and Rhizomucor.

### EORTC Criteria and clinical management

According to the 2019 European Organisation for Research and Treatment of Cancer/Mycoses Study Group (EORTC/MSG) criteria, cases of mucormycosis were classified as proven, probable or putative (16). Following guidelines on the diagnosis and management of mucormycosis from the European Confederation of Medical Mycology (ECMM) and the Mycoses Study Group Education & Research Consortium (MSG ERC), diabetes, burns and trauma were added as risk factors for probable mucormycosis so as not to limit it to immunocompromised patients (18). These criteria were applied whether reported or not in medical observations, and whether the glucose threshold was higher than 1.26 mg per litre of blood on two distinct samples for diabetes. Putative cases were defined as cases with at least one positive PCR result, regardless of the sample site, and without presenting EORTC criteria required for classification as proven or probable. Another classification (EORTC+PCR) was created, considering PCR results in addition to the 2019 EORTC criteria.

### Objectives of the study

The first aim of our study was to provide an epidemiological description of our cohort of patients who tested positive for Mucorales and to assess populations at risk of developing mucormycosis within the cohort. For the analysis, cases of proven or probable mucormycosis were separated from cases of possible mucormycosis, as the former involved treatment initiation. The second aim was to confirm factors associated with in-hospital mortality. The third aim was to assess the diagnostic difficulty of mucormycosis. The diagnoses of three blinded experts were recorded and compared with the EORTC criteria.

### Statistical methods

First, we conducted a multiple correspondence analysis (MCA) to explore the underlying structure of the variables in our dataset. The EORTC classification was included as an additional variable. Multiple imputation was performed using the MCA method to account for missing data while retaining all individuals in the analysis (19,20).

Univariate analyses were conducted for the first two objectives to identify risk factors associated with a proven, probable or possible diagnosis of mucormycosis, and to assess risk factors for in-hospital mortality. Fisher’s exact test was used for categorical variables and the Wilcoxon–Mann–Whitney test for continuous variables. All factors associated with in-hospital mortality that reached a 10% significance level in the univariate analyses were selected as candidate variables for a multivariable logistic regression model. Variables with a high proportion of missing data (n > 10) were excluded, as were those related to length of hospitalisation and time spent in intensive care. An automatic variable selection process using the stepwise method was then applied. To address potential estimation instability due to small sample sizes, Firth’s correction was implemented.

We used the weighted Cohen’s Kappa coefficient to assess the level of consistency in infection diagnoses (proven, probable or putative) between three expert physicians (infectious diseases specialists and microbiologists) and the EORTC criteria. Given that there were three raters, Krippendorff’s alpha was also employed, as this accommodates more than two raters (21).

For all analyses, two-sided p-values < 0.05 were considered statistically significant. MCA was performed using R software with the FactoMineR package (22), while all other statistical analyses were conducted using SAS (version 9.4, SAS Institute, Cary, NC, USA).

## RESULTS

### Patient’s characteristics

#### Clinical characteristics

A total of 85 patient were included (Table 1). The median age was 40 years old, with slightly more males than females. Nine patients had uncontrolled diabetes (17.3%). Eighteen patients presented with HM (23.4%), including eight leukemias. Eleven patients had a history of bone marrow transplantation and eight had a history of solid organ transplantation with immunosuppressive therapy. Eight patients were receiving chemotherapy for active solid cancer. Neutropenia was observed in 17 cases (26.6%).

**Table 1.** Characteristics of patients with mucormycosis (n=85)

|  | All (n=85) |  |
| --- | --- | --- |
|  | n | % |
| **Sex (% of man)** | 47 | 55.3 |
| **Age†** | 40.5 (28.0) | 12-42-66 |
| **ICU (nmiss=7)** | 19 | 24.4 |
| **Hospitalized patients (nmiss=6)** | 60 | 76.0 |
| Length of stay (in days) † (nmiss=2) | 18.8 (20.4) | 3-10-26 |
| **Inhospital death (nmiss=11)** | 10 | 13.5 |
| **Infection site (nmiss=16)** |  |  |
| Cutaneomucosal | 11 | 15.9 |
| Pulmonary | 41 | 59.4 |
| ENT | 11 | 15.9 |
| Other sites | 6 | 8.7 |
| **Uncontrolled diabetes (nmiss=33)** | 9 | 17.3 |
| **Neutropenia (nmiss=21)** |  |  |
| <1.5 G/l | 17 | 26.6 |
| ≥1.5 G/l | 47 | 73.4 |
| **Haematological malignancy (nmiss=8)** | 18 | 23.4 |
| **Transplantation (nmiss=10)** | 19 | 25.3 |
| Bone marrow transplantation | 11 | 57.9 |
| Solid organ transplantation | 8 | 42.1 |
| **Solid organ cancer (nmiss=10)** | 17 | 22.7 |
| Active Cancer (nmiss=1) | 8 | 50.0 |
| **Traumatic injury (nmiss=9)** | 3 | 3.9 |
| **Clinical symptoms** |  |  |
| Neurological symptoms (nmiss=15) | 8 | 11.4 |
| Pneumological symptoms (nmiss=10) | 39 | 52.0 |
| Ears, nose and throat symptoms (nmiss=14) | 14 | 19.7 |
| Cutaneous symptoms (nmiss=11) | 14 | 18.9 |
| Systemic symptoms (nmiss=14) | 21 | 29.6 |
| **Identified species (nmiss=29)** |  |  |
| Rhizomucor *spp.* | 2 | 3.6 |
| Rhizopus *spp.* | 30 | 53.6 |
| Mucor *spp.* | 22 | 39.3 |
| Absidia *spp.* | 2 | 3.6 |
| **Another fungal positive sample (nmiss=22)** | 31 | 49.2 |
| **Culture (nmiss=15)** |  |  |
| ENT | 6 | 8.6 |
| Biopsy | 5 | 83.3 |
| Other sample | 1 | 16.7 |
| Deep respiratory sample | 7 | 10.0 |
| Upper respiratory tract biopsy | 1 | 14.3 |
| Lung biopsy | 1 | 14.3 |
| BAL | 5 | 71.4 |
| Superficial respiratory sample | 33 | 47.1 |
| Bronchial aspiration | 5 | 15.2 |
| Sputum | 28 | 84.8 |
| Others site sample | 24 | 34.3 |
| **Positive PCR** |  |  |
| Blood (nmiss=71) | 5 | 35.7 |
| ENT (nmiss=74) | 7 | 63.6 |
| Biopsy (nmiss=76) | 6 | 66.7 |
| Other sample (nmiss=82) | 2 | 66.7 |
| BAL (nmiss=72) | 9 | 69.2 |
| Superficial respiratory sample (nmiss=74) | 9 | 81.8 |
| Bronchial aspiration (nmiss=78) | 4 | 57.1 |
| Sputum (nmiss=79) | 5 | 83.3 |
| Pleural puncture (nmiss=81) | 1 | 25.0 |
| Cutaneomucosal swab (nmiss=79) | 4 | 66.7 |
| **CT Scanner** |  |  |
| Compatible image (nmiss=40) | 31 | 68.9 |
| Typical image (nmiss=46) | 6 | 15.4 |
| **Surgical procedure performed (nmiss=21)** | 10 | 15.6 |
| **Time between sampling and diagnosis\*** |  |  |
| Culture (nmiss=68) | 11.6 (15.1) | 5-7-9 |
| PCR (nmiss=72) | 5.2 (2.3) | 4-5-7 |
| Histology analysis (nmiss=68) | 23.1 (20.5) | 9-20-28 |
| **Team in charge diagnosis (nmiss=12)** |  |  |
| Proven / Probable | 22 | 30.1 |
| Putative | 51 | 69.9 |
| **Diagnosis with EORTC criteria (nmiss=12)** |  |  |
| Proven / Probable | 11 | 15.1 |
| Putative | 62 | 84.9 |
| **Diagnosis with EORTC + PCR criteria (nmiss=12)** |  |  |
| Proven / Probable | 13 | 17.8 |
| Putative | 60 | 82.2 |

\*Time between sampling and physician assistant taking the microbiological diagnosis in account. ICU: Intensive Care Unit, BAL: Bronchoalveolar lavage, ENT: Ears Nose and Throat, PCR test: Polymerase Chain Reaction test. †: Mean (standard deviation), Q1-Median-Q3

Forty-one (59.4%) and eleven (15.9%) patients were treated for lung and sinus infections, respectively. Nineteen patients were in the intensive care unit (24.4%), with a median length of hospitalisation of 14.2 days. The medical team in charge suspected 22 (30.1%) cases of mucormycosis and started antifungal therapy. Eleven patients (15.1%) had proven or probable mucormycosis according to the EORTC criteria. Ten patients (15.6%) underwent a surgical procedure. Death was reported in ten cases (13.5%), three of which were classified as proven or probable.

#### Laboratory findings

Among the 85 identified patients, 26 (30.5%) had positive Mucorales PCR only, 59 (69.4%) had positive cultures only and 7 (8.2%) had both. Positive cultures on superficial respiratory samples (n=33) were the most reported. For other aera, positive culture were most often reported on biopsy (83.3) for the 6 ear-nose-throat (ENT) samples and on Brochoalveolar lavage (BAL) (71.4%) for the 7 deep respiratory samples. For qPCR tests, 9 positive biopsy PCR assays and 9 BAL positive PCR assays were reported but with less superficial respiratory samples (9). Other Fungal and viral PCR or culture were respectively reported in 31 cases and 11 cases (Table 1).

### Underlying structure of the clinical and laborary characteristics

The first two components of the MCA explained 34% of the total variance, indicating a moderate representation of the structure of the dataset (Figure 2). The graphical representation of the MCA showed clustering of variables associated with disease severity on the right side of the first axis, such as ICU admission and death, suggesting a strong correlation between these factors. Conversely, variables associated with lower disease severity were projected on the left side of the first axis. The EORTC classification, included as an illustrative variable, was positioned close to the clinical severity indicators, highlighting its potential association with these variables.

**Figure 2.**
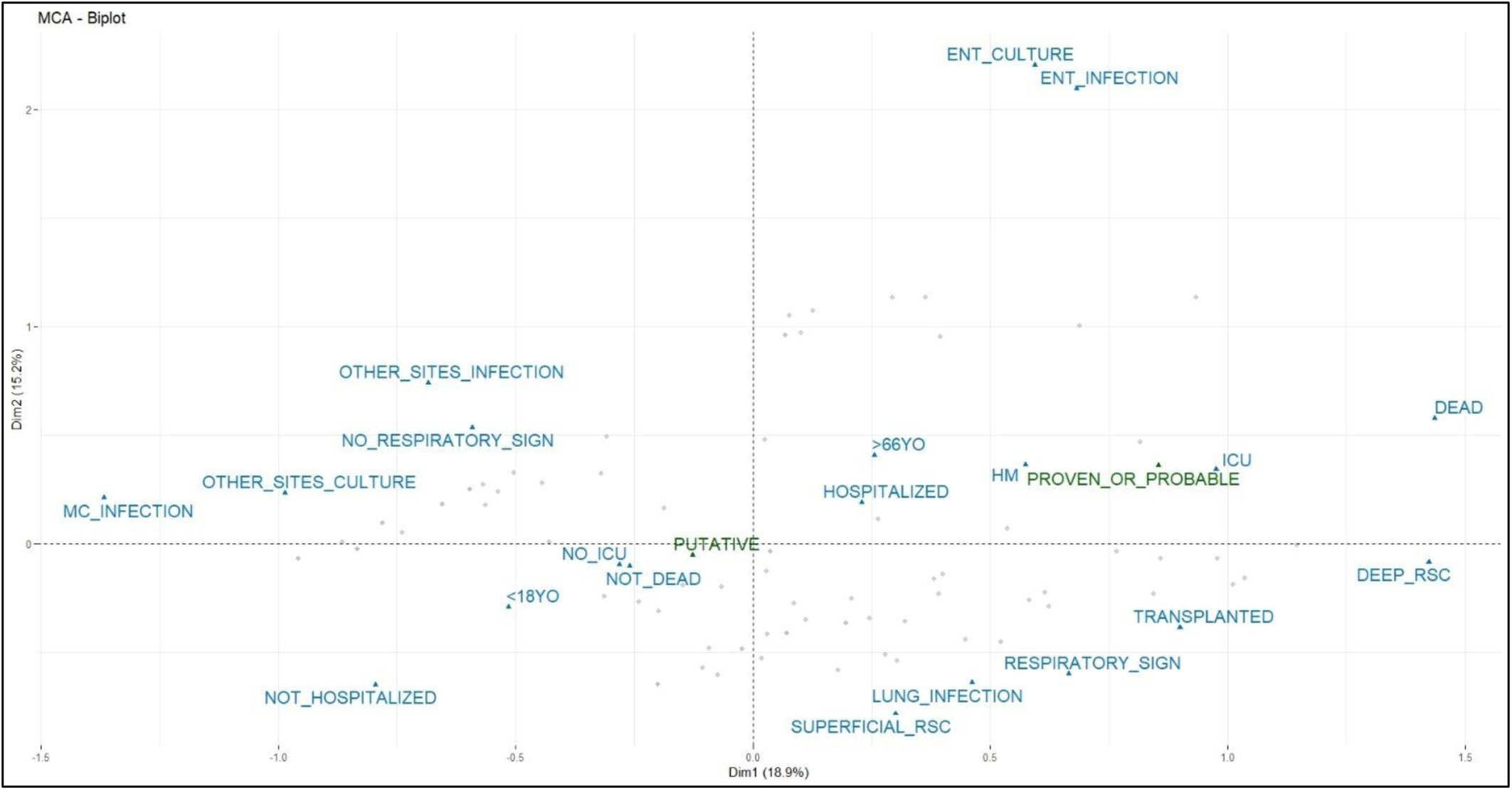
Multiple Correspondence Analysis (MCA) (n=85) Variables: Sex, resuscitation, hospitalisation, infection site, age class, death, haemopathy, transplant, cancer, culture, pneumological signs, EORTC 2019 class (illustrative variable). The first two factorial axes explained 34% of the total variance. To keep all individuals in the analysis, missing data were imputed by multiple ACM. MC infection: Mucocutaneous infection, ENT infection: Ear, nose and throat infection, HM = Haematological malignancy, Deep RSC: Deep respiratory samples culture, Superficial RSC: Superficial respiratory samples culture,

### Risk Factors of mucormycoses diagnosis when having a sample positive

Compared to possible mucormycosis patients, proven or probable mucormycosis patients were more likely to be hospitalised in intensive care (p=0.011) and to have a longer hospital stay (30.5 days, p=0.011). Eight out of eleven cases had a history of transplantation (p<0.001), and the use of surgery was significantly increased (p<0.001). Although not statistically significant, the proven or probable mucormycosis group had a reported death rate of 30%, compared to 8% (p=0.08). There were also more ENT positive samples in the proven or probable mucormycosis group than in the possible mucormycosis group (p=0.05) (Table 2).

**Table 2.** Characteristics of mucormycosis patients following EORTC criteria (n=73)

|  | Proven or Probable case(n=11) |  | Putative case (n=62) |  | p-value† |
| --- | --- | --- | --- | --- | --- |
|  | n | % | n | % |  |
| **Sex (% of man)** | 5 | 45.5 | 34 | 54.8 | 0.745 |
| **Age‡** | 35.8 (27.5) | 12-25-66 | 37.3 (28.1) | 7-36-62 | 0.957 |
| **ICU** | 6 | 54.5 | 10 | 16.1 | **0.011** |
| **Hospitalized patient** | 10 | 90.9 | 45 | 72.6 | 0.273 |
| Length of stay (in days) ‡ (nmiss=2) | 30.5 (22.0) | 10-34-51 | 16.8 (20.0) | 2-8-23 | 0.080 |
| **Inhospital death (nmiss=3)** | 3 | 30.0 | 5 | 8.3 | 0.081 |
| **Infection site (nmiss=8)** |  |  |  |  |  |
| Cutaneomucosal | 0 | 0.0 | 10 | 18.5 | 0.190 |
| Pulmonary | 6 | 54.5 | 34 | 63.0 | 0.737 |
| ENT | 4 | 36.4 | 7 | 13.0 | 0.080 |
| Other sites | 1 | 9.1 | 3 | 5.6 | 0.533 |
| **Uncontrolled diabetes (nmiss=23)** | 2 | 22.2 | 7 | 17.1 | 0.657 |
| **Neutropenia (nmiss=13)** |  |  |  |  |  |
| <1.5 G/l | 4 | 36.4 | 9 | 18.4 | 0.231 |
| ≥1.5 G/l | 7 | 63.6 | 40 | 81.6 | 0.231 |
| **Haematological malignancy (nmiss=2)** | 4 | 36.4 | 11 | 18.3 | 0.228 |
| **Transplantation (nmiss=3)** | 8 | 72.7 | 9 | 15.3 | **<.001** |
| Bone marrow transplantation | 6 | 75.0 | 3 | 33.3 | 0.153 |
| Solid organ transplantation | 2 | 25.0 | 6 | 66.7 | 0.153 |
| **Solid organ cancer (nmiss=3)** | 2 | 18.2 | 13 | 22.0 | 1.000 |
| Active Cancer (nmiss=1) | 0 | 0.0 | 6 | 50.0 | 0.473 |
| **Clinical respiratory signs (nmiss=2)** | 6 | 54.5 | 30 | 50.0 | 1.000 |
| **Culture (nmiss=12)** |  |  |  |  |  |
| ENT | 3 | 30.0 | 3 | 5.9 | 0.050 |
| Biopsy | 3 | 100.0 | 2 | 66.7 |  |
| Other sample | 0 | 0.0 | 1 | 33.3 |  |
| Deep respiratory sample | 2 | 20.0 | 5 | 9.8 | 0.322 |
| Upper respiratory tract biopsy | 1 | 50.0 | 0 | 0.0 |  |
| Lung biopsy | 1 | 50.0 | 0 | 0.0 |  |
| BAL | 0 | 0.0 | 5 | 100.0 |  |
| Superficial respiratory sample | 4 | 40.0 | 29 | 56.9 | 0.490 |
| Bronchial aspiration | 0 | 0.0 | 5 | 17.2 |  |
| Sputum | 4 | 100 | 24 | 82.7 |  |
| Others site sample | 1 | 10.0 | 14 | 27.5 | 0.426 |
| **Positive PCR** |  |  |  |  |  |
| Blood (nmiss=62) | 1 | 25.0 | 2 | 28.6 | 1.000 |
| ENT (nmiss=62) | 2 | 66.7 | 5 | 62.5 | 1.000 |
| Biopsy (nmiss=64) | 2 | 66.7 | 4 | 66.7 | 1.000 |
| Other sample (nmiss=70) | 1 | 100.0 | 1 | 50.0 | 1.000 |
| BAL (nmiss=62) | 3 | 100.0 | 5 | 62.5 | 0.491 |
| Superficial respiratory sample (nmiss=62) | 3 | 75.0 | 6 | 85.7 | 1.000 |
| Bronchial aspiration (nmiss=66) | 2 | 66.7 | 2 | 50.0 | 1.000 |
| Sputum (nmiss=67) | 1 | 50.0 | 4 | 100.0 | 0.333 |
| Pleural puncture (nmiss=70) | 1 | 50.0 | 0 | 0.0 | 1.000 |
| Cutaneomucosal swab (nmiss=67) | 1 | 50.0 | 3 | 75.0 | 1.000 |
| **Other fungal positive sample (nmiss=11)** | 5 | 50.0 | 25 | 48.1 | 1.000 |
| **Surgical procedure performed (nmiss=11)** | 6 | 66.7 | 4 | 7.5 | **<.001** |

†: Fisher's exact test for qualitative variables; Wilcoxon Mann Whitney test for quantitative variables.
‡: Mean (standard deviation), Q1-Median-Q3

### Risk Factors of in-hospital mortality, univariate analysis

While assessing risk factors for death in our population, we found that differences in ICU admission (p < 0.001), length of hospital stay (mean length of stay in days: 26.0 vs. 16.3, p = 0.012) and neutropenia were significant (p < 0.03). (Table 3). Four out of five patients with uncontrolled diabetes died (p = 0.028). Those who died had more positive deep lung samples (p = 0.022) and were more easily treated by the medical team in charge (p < 0.01). Although no significant difference was observed, sinus infection tended to be more prevalent in the deceased group (p = 0.054). A diagnosis of proven or probable mucormycosis following EORTC criteria was not associated with death in our population (p = 0.081), but significance was reached when patients were classified according to the modified EORTC + PCR criteria, taking PCR into account (p = 0.003) (Table 3).

**Table 3.** Characteristics of mucormycosis patients according to death (n=74)

|  | Death (n=10) |  | No death (n=64) |  | p-value† |
| --- | --- | --- | --- | --- | --- |
|  | n | % | n | % |  |
| **Sex (% of man)** | 5 | 50.0 | 35 | 54.7 | 1.000 |
| **Age‡** | 44.3 (26.6) | 20-47.5-66 | 37.1 (27.8) | 7.5-35.5-62 | 0.957 |
| **ICU** | 8 | 80.0 | 8 | 12.5 | **<.001** |
| **Hospitalized patient** | 10 | 100.0 | 45 | 70.3 | 0.056 |
| Length of stay (in days) ‡ (nmiss=2) | 26.0 (11.6) | 23-24-30 | 16.3 (21.1) | 2-8-19 | **0.012** |
| **Infection site (nmiss=8)** |  |  |  |  |  |
| Cutaneomucosal | 0 | 0.0 | 11 | 19.6 | 0.192 |
| Pulmonary | 5 | 50.0 | 34 | 60.7 | 0.729 |
| ENT | 4 | 40.0 | 7 | 12.5 | 0.054 |
| Other sites | 1 | 10.0 | 4 | 7.1 | 0.573 |
| **Uncontrolled diabetes (nmiss=25)** | 4 | 50.0 | 5 | 12.2 | **0.028** |
| **Neutropenia (nmiss=14)** |  |  |  |  |  |
| <1.5 G/l | 5 | 50.0 | 8 | 16.0 | **0.031** |
| ≥1.5 G/l | 5 | 50.0 | 42 | 84.0 | **0.031** |
| **Haematological malignancy (nmiss=2)** | 3 | 30.0 | 11 | 17.7 | 0.397 |
| **Transplantation (nmiss=3)** | 3 | 30.0 | 14 | 23.0 | 0.694 |
| Bone marrow transplantation | 0 | 0.0 | 9 | 64.3 | 0.082 |
| Solid organ transplantation | 3 | 100.0 | 5 | 35.7 | 0.082 |
| **Solid organ cancer (nmiss=4)** | 4 | 40.0 | 12 | 20.0 | 0.221 |
| Active Cancer (nmiss=1) | 2 | 50.0 | 5 | 45.5 | 1.000 |
| **Clinical respiratory signs (nmiss=3)** | 7 | 70.0 | 29 | 47.5 | 0.307 |
| **Culture (nmiss=12)** |  |  |  |  |  |
| ENT | 1 | 12.5 | 5 | 8.9 | 0.567 |
| Biopsy | 1 | 100.0 | 4 | 80.0 |  |
| Other sample | 0 | 0.0 | 1 | 20.0 |  |
| Deep respiratory sample | 3 | 37.5 | 3 | 5.4 | **0.022** |
| Upper respiratory tract biopsy | 1 | 33.3 | 0 | 0.0 |  |
| Lung biopsy | 1 | 33.3 | 0 | 0.0 |  |
| BAL | 1 | 33.3 | 3 | 100.0 |  |
| Superficial respiratory sample | 2 | 25.0 | 31 | 55.4 | 0.142 |
| Bronchial aspiration | 2 | 100.0 | 3 | 9.7 |  |
| Sputum | 0 | 0.0 | 14 | 90.4 |  |
| Others site sample | 2 | 25.0 | 17 | 30.4 | 1.000 |
| **Positive PCR** |  |  |  |  |  |
| Blood (nmiss=63) | 2 | 66.7 | 2 | 25.0 | 0.491 |
| ENT (nmiss=63) | 4 | 100.0 | 3 | 42.9 | 0.194 |
| Biopsy (nmiss=65) | 3 | 100.0 | 3 | 50.0 | 0.464 |
| Other sample (nmiss=71) | 2 | 100.0 | 0 | 0.0 | 0.333 |
| BAL (nmiss=65) | 3 | 100.0 | 3 | 50.0 | 0.464 |
| Superficial respiratory sample (nmiss=64) | 1 | 100.0 | 7 | 77.8 | 1.000 |
| Bronchial aspiration (nmiss=68) | 1 | 100.0 | 2 | 40.0 | 1.000 |
| Sputum (nmiss=68) | - | - | 5 | 83.3 | - |
| Pleural puncture (nmiss=72) | - | - | 0 | 0.0 | - |
| Cutaneomucosal Swab (nmiss=69) | 2 | 100.0 | 1 | 33.3 | 0.400 |
| **Team in charge diagnosis (nmiss=4)** |  |  |  |  |  |
| Proven / Probable | 8 | 100.0 | 12 | 19.4 | **<.001** |
| Putative | 0 | 0.0 | 50 | 80.6 | **<.001** |
| **Diagnosis with EORTC criteria (nmiss=4)** |  |  |  |  |  |
| Proven / Probable | 3 | 37.5 | 7 | 11.3 | 0.081 |
| Putative | 5 | 62.5 | 55 | 88.7 | 0.081 |
| **Diagnosis with EORTC +PCR criteria (nmiss=4)** |  |  |  |  |  |
| Proven / Probable | 5 | 62.5 | 7 | 11.3 | **0.003** |
| Putative | 3 | 37.5 | 55 | 88.7 | **0.003** |
| **Other fungal positive sample (nmiss=14)** | 5 | 62.5 | 25 | 48.1 | 0.706 |
| **Surgical procedure performed (nmiss=13)** | 2 | 20.0 | 7 | 13.7 | 0.633 |

†: Fisher's exact test for qualitative variables; Wilcoxon Mann Whitney test for quantitative variables. ‡: Mean (standard deviation), Q1-Median-Q3

**Table 4.** Factors associated with death – Multivariable logistic regression (n=74)

| Variable | Odds ratio | 95%CI | p-value |
| --- | --- | --- | --- |
| Positive culture deep respiratory sample | 16.63 | 1.67 165.58 | 0.017 |
| Proven or Probable with EORTC + PCR criteria | 12.37 | 1.59 96.30 | 0.016 |

**Table 5.** Script concordance tests (n=70)

| Weighted Kappa coefficient<br>[95% confidence interval] |  | Expert clinicians |  | EORTC criteria |
| --- | --- | --- | --- | --- |
|  |  | n°2 | n°3 |  |
| Expert clinicians | n°1 | 0.403 [0.204 – 0.602] | 0.346 [0.147 – 0.544] | 0.204 [0.054 – 0.354] |
|  | n°2 |  | 0.293 [0.141 – 0.444] | 0.125 [0.039 – 0.211] |
|  | n°3 |  |  | 0.508 [0.298 – 0.718] |

Krippendorff's alpha was estimated at 0.243, indicating low overall agreement between the three doctors and the clinical opinion (n=70).

### Risk Factors of in-hospital mortality, Mutivariate analysis

Following stepwise selection, two risk factors were found to be significantly associated with in-hospital mortality in the multivariable model: a positive deep respiratory culture test result (odds ratio (OR) = 16.6, p = 0.017) and a proven or probable diagnosis according to the EORTC+PCR criteria (OR = 12.4, p = 0.016).

### Consistency in the diagnoses of mucormycoses

Comparing mucormycosis diagnoses using the EORTC and EORTC + PCR criteria showed a high agreement, with a Kappa coefficient of 0.90 (95% CI: [0.77–1.00]). A total of 97.3% of cases were classified consistently using both sets of criteria, with only two cases (2.7%) showing discrepancies.

By contrast, the inter-observer agreement among the three blinded experts was lower, with Kappa coefficients ranging from 0.29 to 0.40 indicating a weak to moderate level of agreement. The Krippendorff’s alpha coefficient was 0.32 overall, reflecting low to moderate consistency between the three experts.

Agreement varied widely when comparing experts’ diagnoses with the EORTC criteria, with Kappa coefficients ranging from 0.13 to 0.51. The Krippendorff’s alpha of 0.24 overall suggests weak agreement between the physicians and the EORTC 2019 classification. These findings reveal substantial variability in clinical diagnoses among the experts.

## DISCUSSION

Due to the lack of data on these rare but severe fungal diseases, we strongly believe that our bicentric retrospective study could be a relevant addition to recent large-scale studies focusing on mucormycosis (3,5,8). Our laboratory provide diagnostic assays over clinical specimens obtained from a wide range of patients (intensive care, haematological intensive care, general medical units). Clinical analysis combined with modern laboratory testing and mycological expertise could help clinicians to differentiate between Mucorales colonisation and infection, thus improving clinical management.

Several cases in the literature describe Mucorales colonisation in various anatomical aera, in regard to the presence of this fungus in human environments (23–26). One Iranian study found this rate to be as high as 12% of patients for which cutaneous samples had been collected (24). In addition, fungal colonisation has been described as a risk factor for invasive infections in transplant patients (27). However, some Mucorales species have never been reported to cause invasive infections, underlining the importance of species identification when possible (23).

Most cases of misdiagnoses either result in mistakenly treating mucormycosis based on explosive clinical symptoms or neglecting the diagnosis of insidious forms, both of which may be the consequence of poorly sensitive microbiological diagnostic methods (28–30). With improved diagnostic techniques, situations of microbiological deficiency are tending to diminish in favour of overtreatment of situations in which a fungus is identified without information on its role in existing symptoms (11,31,32). As this rapidly progressing infection continues to have a negative prognosis, it appears appropriate to treat patients until a clear distinction is made between colonization and infection.

This study was launched in response to clinicians’ question about what to do when facing isolated mucormycetes positive samples especially in vulnerable patient. It is the only study to our knowledge that focus on mucor positive sample instead of being diagnosed with mucormycosis as inclusion criteria. Both infections and colonizations are presented, to highlight the differences between these two populations. The large number of positive cultures on superficial samples (33 patients) and the small number of proven and probable cases (11 cases) in our cohort raises questions about the prevalence of Mucorales colonization.

In our study, 22.2% of patients with proven or probable mucormycosis presented with uncontrolled diabetes (N= 2). This is much lower than in Indian studies, which report uncontrolled diabetes in 56.8% to 73.6% of patients (33,34). However, our results are consistent with the prevalence of diabetes in the European Confederation of Medical Mycology (17%) and the Retrozygo study (23%). Therefore, we could speculate about a correlation between the mortality rate of mucormycosis and the rate of imbalance of diabetes mellitus (35). We could not find evidence of this in our statistical analysis, but some studies have shown that the key to infection is the effect of an acidic pH on the potential of fungal pathogens. An acidic pH is often found in patients with very imbalanced diabetes due to ketoacidosis. Thus, in this population, the key question about mucormycosis infection and mortality risk remains unanswered, and, in light of our analyses, we strongly believe that a maximalist and preventive approach is necessary with this group of patients.

There were no significant differences in cultures or PCR results between patients with proven or probable mucormycosis for every specimen types, whereas another study found significant differences in favour of PCR use in managing mucormycosis in patients with haematological malignancies (36). One hypothesis is that our cohort included four cases of rhino-orbito-cerebral mucormycosis (ROCM). Providing a better sample with surgery for ROCM may increase culture sensitivity while appears to decrease PCR sensitivity (10). Furthermore, Very few serum specimens were available while studies on mucor PCRs are mainly targeting serum PCRs (3,8,14,36,37). Few data have been reported recently specifically for ROCM, or for PCR on other superficial or deep samples with reasonable sensitivity and good specificity (>97%) (10,12).

In addition, we compared the time between sampling and clinicians taking the microbiological diagnosis into account, and between culture and PCR for all samples combined. The median was 7 days and 5 days respectively. This is consistent with, but less than, a recent publication which found that serum PCR took about 4 days earlier (36). These data suggest that the precise role of PCR remains to be defined, but that it can be an asset in optimising diagnostic time and therapeutic strategy (14).

In terms of in-hospital mortality, principal component analysis revealed a homogeneous group of patients with positive deep respiratory swabs, haematology, ICU admission and a higher mortality rate. This is consistent with previous studies despite the severity of our patients’ underlying conditions (4,7,9,38). Patients who died were significantly more neutropenic. One of the EORTC’s diagnostic criteria is profound neutropenia (defined as <0.5 × 10^9^ neutrophils/L for >10 days) (16). However, very few patients in our neutropenic group (<1.5 × 10⁹ neutrophils/L in the month of mucormycosis diagnosis) completely met the EORTC’s criteria for neutropenia. This suggests the need for caution when diagnosing mucormycosis and managing immunocompromised patients, regardless of the severity of the neutropenia.

However, our study has a number of design limitations. First, many missing data were not reported at the time of medical management, which affects the logistic regression estimates, as shown by the size of the confidence intervals. In addition, the small size of our population exposes the study to lack of power. Secondly, only PCR-positive cases were reported and not the number of cases tested by PCR or the number of PCR tests performed by patients, which would have been relevant. Finally, the criterion of in-hospital mortality suffers from a degree of imprecision when it comes to the actual outcome of patients, although it does reflect the severity of contemporary infection.

The low level of agreement between expert physicians in our study also highlights the diagnostic difficulties faced by clinicians in these life-threatening infections. With this in mind, by including positive PCRs in the EORTC recommendations, we were able to move 2 patients into the proven/probable category. This was reflected in the emergence of a significant difference in in-hospital mortality in the proven/probable MCM group (EORTC+PCR, Table 3). This raises the question of reconsidering the role of the deep PCR technique in the diagnostic approach (10,12,36), provided that the same rigor as for Aspergillus PCRs is applied (i.e. at least 2 BAL or blood PCRs, or one of each) in the EORTC guidelines (10,16).

## CONCLUSION

Mucormycosis remains a rare but serious fungal infection affecting mainly immunocompromised patients. Despite advances in diagnostic tools and treatment modalities, high in-hospital mortality rates are reported, particularly in cases of pulmonary or disseminated infection, highlighting the importance of rapid diagnostic methods. Our study provides valuable insights into the epidemiological and clinical characteristics of patients with positive mucormycosis specimens in a French cohort.

We observed that proven or probable mucormycosis was more frequently associated with intensive care admission, longer hospital stay and higher in-hospital mortality. Factors such as uncontrolled diabetes, haematological malignancies and solid organ transplantation emerged as significant risk factors, highlighting the need for early detection and tailored management strategies. The role of PCR in diagnostic accuracy appears to be relevant, particularly when combined with traditional EORTC criteria, suggesting that diagnostic guidelines may need to be refined to effectively incorporate these tools. However, our study is limited by its retrospective design, small sample size, and missing data, which may affect the precision of our findings. As recently suggested, prospective studies with larger cohorts are needed to validate our findings and further elucidate risk factors, diagnostic challenges, and optimal treatment approaches for mucormycosis. Filling these gaps will be critical to improving outcomes for this often overlooked but life-threatening infection.

## Data Availability

clinical and microbiological data

## DECLARATIONS

### Ethics approval and consent to participate

This study complies with the General Data Protection Regulation and with obligations of the Commission Nationale de l’Informatique et des Libertés, MR004 reference methodology according to French law (MR004-PADS23-94). This study has been registered and can be consulted on the Health Data Hub’s MR 004 public directory (n°16210117).

Data collection was carried out in accordance with the legal procedures imposed by each centre: Pseudonymisation was used for data collection. No information other than those collected at the time of medical management was considered. For the Assistance Publique des Hôpitaux de Marseille (APHM) centres, the non-objection of patients was sought by consulting the hospital register of patient refusals. Agreements of the chiefs of department in which the patients were hospitalised were also requested and obtained. For the cancer centre, an official agreement was signed and is available in supplementary data (RCAPHM24_0149_A_0001). Written information to patient was sent with a one-month objection period.

This study was approved by the APHM internal ethic Review Board in November 2023 (Decision No. CSE23-49).

## Availability of data and materials

The datasets used and/or analysed during the current study are available from the corresponding author on reasonable request.

## Competing interests

The authors have no conflicts of interest to declare. Funding sources had no role in the design and conduct of the study; collection, management, analysis, and interpretation of the data; and preparation, review, or approval of the manuscript.

## Funding

No funding was involved.

## Author contributions

Conceived and designed the experiments: NS JS PD HBeu JCL SR. Contributed materials/analysis tools: LD SC PV HBe HBeu. Analyzed the data: LD SC PD HBeu HBe PV NC JS. Wrote the paper: PD HBeu JS NC JCL. Revised the draft: HBeu, JS, HBe, PV JHCL NC SR PD.

## Acknowledgments

We strongly thank all administration staff from Assistance Publique Hôpitaux de Marseille and Institut Paoli Calmette.

## Notes

### Competing Interest Statement

The authors have declared no competing interest.

### Funding Statement

no fundings

### Author Declarations

The Ethics and Scientific Committee (CSE) of the AP-HM met on 21/11/2023. It examined your project "Retrospective study of cases of mucormycosis diagnosed at the APHM between 2017 and 2022," with particular attention paid to: - The scientific justification for the objective - The feasibility of the study - The admissibility of patient information - Ethical aspects The assessment of these various items led the CSE to issue a favourable opinion on the project "Retrospective study of cases of mucormycosis diagnosed at the APHM between 2017 and 2022." You will also find attached an information notice based on the details of your project, which you can use as a basis for applying the instructions on respecting the rights of individuals that have been given to you elsewhere (by the PADS and/or the medical evaluation service, depending on your contact person). Prof. Jean-Robert Harle Chair of the CSE AP-HM

